# Deep Learning Model Utilization for Mortality Prediction in Mechanically Ventilated ICU Patients

**DOI:** 10.1101/2024.03.20.24304653

**Authors:** Negin Ashrafi, Yiming Liu, Xin Xu, Yingqi Wang, Zhiyuan Zhao, Maryam Pishgar

## Abstract

**Background:** The requirement for mechanical ventilation has increased in recent years. Patients in the intensive care unit (ICU) who undergo mechanical ventilation often experience serious illness, contributing to a high risk of mortality. Predicting mortality for mechanically ventilated ICU patients helps physicians implement targeted treatments to mitigate risk.

**Methods:** We extracted medical information of patients with invasive mechanical ventilation during ICU admission from the Medical Information Mart for Intensive Care III (MIMIC-III) dataset. This information includes demographics, disease severity, diagnosis, and laboratory test results. Patients who met the inclusion criteria were randomly divided into the training set (n=11,549, 70%), the test set (n=2,475, 15%), and the validation set (n=2,475, 15%). The Synthetic Minority Over-sampling Technique (SMOTE) was utilized to resolve the imbalanced dataset. After literature research, clinical expertise and an ablation study, we selected 12 variables which is fewer than the 66 features in the best existing literature. We proposed a deep learning model to predict the ICU mortality of mechanically ventilated patients, and established 7 baseline machine learning (ML) models for comparison, including K-nearest Neighbors (KNN), Logistic Regression, Decision Tree, Random Forest, Bagging, XGBoost, and Support Vector Machine (SVM). Area under the Receiver Operating Characteristic Curve (AUROC) was used as an evaluation metric for model performance.

**Results:** Using 16,499 mechanically ventilated patients from the MIMIC-III database, the Neural Network model outperformed existing literature by 7.06%. It achieved an AUROC score of 0.879 (95% Confidence Interval (CI) [0.861-0.896]), an accuracy of 0.859 on the test set, and was well-calibrated with a Brier score of 0.0974, significantly exceeding previous best results.

**Conclusions:** The proposed model demonstrated an exceptional ability to predict ICU mortality among mechanically ventilated patients. The SHAP analysis showed respiratory failure is a significant indicator of mortality prediction compared to other related respiratory dysfunction diseases. We also incorporated mechanical ventilation duration variable for the first time in our prediction model. We observed that patients with higher mortality rates tended to have longer mechanical ventilation times. This highlights the model’s potential in guiding clinical decisions by indicating that longer mechanical ventilation may not necessarily enhance patient survival chances.

## 1. BACKGROUND

An intensive care unit (ICU) is designated for individuals facing severe illnesses or injuries. Most of these patients require assistance from medical equipment, such as mechanical ventilation, to sustain normal bodily functions, and they need to be monitored continuously and intensively [1, 2, 3].

Mechanical ventilation is a crucial life-support method for critically ill patients in the intensive care unit (ICU). In the ICU, more than 25% of patients require mechanical ventilation [4], and approximately 40% of ICU patients in the United States receive invasive mechanical ventilation at any given time [7]. Despite its importance in supporting organ function [6], the use of invasive mechanical ventilation is associated with a high risk of mortality and various complications [5], resulting in notably high mortality rates among patients requiring this intervention [8]. Furthermore, the use of mechanical ventilation contributes to 12% of overall hospital expenditures in the United States [7], highlighting its significant financial impact.

With the growing life expectancies and extended survival times of individuals with chronic conditions, the utilization of mechanical ventilation for artificial support is anticipated to increase [9, 10]. Mechanically ventilated patients usually experience acute respiratory failure or reduced lung function due to an underlying condition, such as pneumonia, sepsis, or heart disease [11, 12, 13]. Alternatively, the need for respiratory assistance may arise from neurological disabilities, disorders of consciousness, or fatigue after significant surgical procedures [14].

In recent years, machine learning algorithms have been widely employed to predict diverse critical health outcomes [15, 16], more specifically those associated with mechanical ventilation [17, 18]. Developing a mortality prediction model for patients with mechanical ventilation may offer valuable support to ICU physicians for timely alerts and informed clinical judgment [19].

Neural network modeling has gained widespread recognition for its effectiveness and it has become a powerful tool for sophisticated modeling in various domains [20]. Neural networks employ a multi-layered structure to autonomously generate distinctive features. Each neuron in the network computes a weighted sum of its inputs, which is then passed through a nonlinear activation function. As a result, neural networks often demonstrate an advantage over traditional machine learning models, such as Logistic Regression, Decision Trees, or SVM, in capturing nonlinearities, particularly when a large amount of data is available [21, 22]. However, the performance advantage of neural networks may vary depending on the size of the dataset.

The primary aim of this research was to establish a deep learning model designed for forecasting the mortality of ICU patients undergoing mechanical ventilation, utilizing comprehensive patient medical history data. Our model achieved higher evaluation performance compared to the best existing literature while using fewer predictive variables.The inclusion of additional variables CHF and respiratory failure significantly enhanced the results of our proposed model. The predictive model was achieved based on guidelines of the Transparent Reporting of Individual Prognostic or Diagnostic Multivariate Predictive Model (TRIPOD) initiative.

## 2. METHODOLOGY

### 2.1. Data Source and Study Design

The Medical Information Mart for Intensive Care (MIMIC-III) database, a comprehensive database containing rich clinical patient data, was used in our study [23]. Specific data, including patients’ clinical physiological parameters and disease diagnosis reports, were extracted from the database to cover specific patient cohorts. We selected the MIMIC-III database because it provides a substantial amount of real-world patient data. This data contributes to a more comprehensive understanding of the research questions and hypotheses. Following the completion of data extraction, necessary data preprocessing was conducted to ensure data quality and alignment with the requirements for model training. The data from the MIMIC-III database provided a crucial foundation for our study, offering robust support for in-depth analysis and model construction, benefiting medical institutions and researchers.

### 2.2. Patient extraction

Our study focused on adult patients who underwent invasive mechanical ventilation during their ICU stay. Figure 1 shows the patient extraction process. First, we selected 61,532 patients with ICU stays records and extracted 28,861 patients whose records indicated a ventilation duration greater than 0. Among those patients, we excluded patients under 18 or over 90 years of age upon ICU admission, as well as patients with missing records of relevant physiological indicators. Initially, we encountered 51 rows with missing values in five features (Minimum PaO2, Maximum PaCO2, Minimum PaCO2, Minimum Lactate, Minimum BUN), representing only 0.036% of our dataset (16,550 patients). Given that this was less than 1% of the data, we removed these rows, assuming it would not affect our results. However, to ensure the robustness of our approach, we also imputed the missing values using the mean of the respective features and found that the model’s accuracy remained consistent, confirming that either approach did not affect the model’s effectiveness. In the end, we extracted a total 16,499 patients who met the established inclusion criteria for the final analyses.

**Figure 1:**
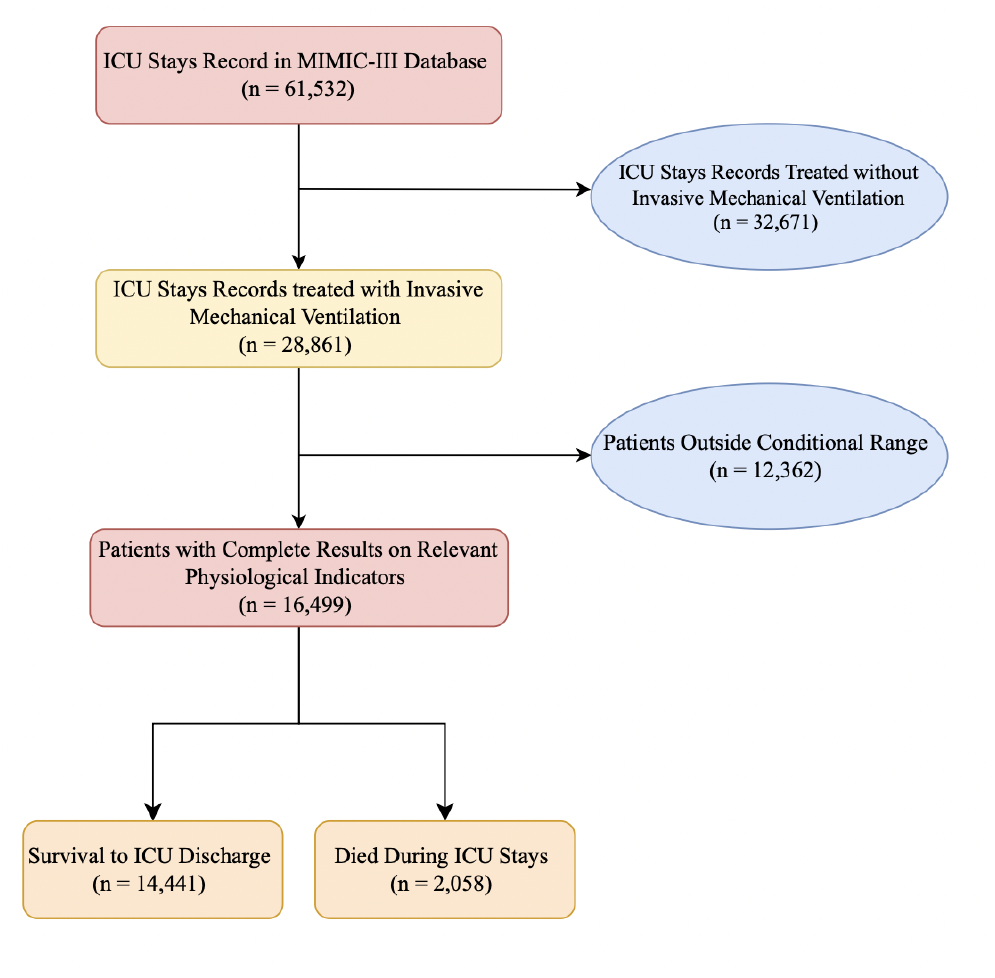
Flow diagram of the selection process of patients.

### 2.3. Statistical analysis between cohorts

The train and validation cohorts were compared using Chi-Square tests and two-sided t-tests with a significance level of P < 0.05 to help determine whether there are significant differences between the training set and the validation set. Chi-Square tests were utilized for comparing categorical variables, while t-tests were employed for continuous variables.

### 2.4. Feature selection

We started with 65 variables based on literature research and expert opinion. Initially, we excluded features with more than 80% missing values. Then, we used the XGBoost model to calculate the importance of the remaining features, excluding those with importance below the threshold.This process left us with 14 key predictors that were chosen for their high importance scores and documented impact on patient outcomes. The subject IDs and ICU stay IDs serve as the unique identifiers for patients and records of ICU admission, respectively. All physiological test indicators and disease diagnoses were referred to ICD-9 codes. Table 1 illustrates the proposed 14 predictors, including: (i) age: patients’ age when entering the ICU; (ii) respiratory dysfunction: all diseases related to ‘respiratory’ in the diagnostic table; (iii) SAPS II: Simplified Acute Physiology Score II; (iv) maximum hemoglobin: patients’ maximum value of blood hemoglobin in the lab events records; (v) minimum lactate: patients’ minimum levels of lactate in the lab events records; (vi) respiratory failure: patients diagnosed with respiratory failure; (vii) minimum BUN: patients’ minimum levels of blood urea nitrogen in the lab events records; (viii) CHF: patients diagnosed with chronic heart failure; (ix) diabetes: patients diagnosed with diabetes; (x) malignancy: patients diagnosed with malignancy; (xi) maximum PaCO2: patients’ maximum levels of partial pressure of carbon dioxide in the arterial blood; (xii) minimum PaCO2: patients’ minimum levels of partial pressure of carbon dioxide in the arterial blood; (xiii) vent duration: the duration of invasive mechanical ventilation; and (xiv) maximum PaO2: patients’ maximum levels of partial pressure of oxygen in the arterial blood.

**Table 1.** Features category table. Demographic: Age. Disease severity: SAPS II. Diagnosis: Respiratory dysfunction, Respiratory failure, CHF, Diabetes, Malignancy. Laboratory results: Maximum hemoglobin (g/dl), Minimum lactate (mmol/L), Minimum BUN (mg/dl), Minimum PaO2(mmHg), Maximum PaCO2(mmHg), Minimum PaCO2(mmHg). Others: Vent Duration (Hour).

| Category | Features | Category | Features |
| --- | --- | --- | --- |
| Demographic | Age (years) | Laboratory results | Maximum hemoglobin (g/dl) |
| Disease severity | SAPS II |  | Minimum lactate (mmol/L) |
| Diagnosis | Respiratory dysfunction |  | Minimum BUN (mg/dl) |
|  | Respiratory failure |  | Minimum PaO<sub>2</sub>(mmHg) |
|  | CHF |  | Maximum PaCO<sub>2</sub>(mmHg) |
|  | Diabetes |  | Minimum PaCO<sub>2</sub>(mmHg) |
|  | Malignancy | Others | Vent Duration (Hour) |

First, we applied the XGBoost model to get the feature importance of these variables and we selected the top 5 important features: age, respiratory dysfunction, SAPS II score, maximum hemoglobin, and minimum lactate. XG-Boost is a powerful ML model which has been widely used for feature selection [19, 24]. This model includes regularization parameters such as ‘gamma’, ‘alpha’, and ‘lambda’ that help prevent overfitting, ensuring that the selected features are not chosen purely based on noise in the data.

After conducting related research and considering expert opinion, we also added other possible influencing factors: malignancy, BUN (Blood urea nitrogen), CHF (Congestive heart failure), diabetes, vent duration, respiratory failure, maximum PaCO2, minimum PaCO2, and maximum PaO2 as our variables. We decided to include these factors because their impact on respiratory health is well-documented and they have the potential to significantly impact outcomes for patients requiring mechanical ventilation.

Malignancies can directly affect the respiratory system, such as lung cancer or metastases to the lungs, leading to compromised lung function. Especially in advanced stages, it can be a crucial factor leading to respiratory failure and the need for mechanical ventilation [25]. BUN is a medical test that measures the amount of urea nitrogen found in blood. High BUN levels can be associated with conditions that may lead to respiratory failure, such as severe infections, sepsis, or organ dysfunction. The decision to use mechanical ventilation is based on a combination of factors, including the underlying condition causing the high BUN level [26]. CHF can cause pulmonary effusion (pulmonary edema), leading to severe breathing difficulties and respiratory failure, thus requiring the use of mechanical ventilation to support breathing [24]. Diabetes can contribute to conditions such as respiratory infections, acute respiratory distress syndrome (ARDS), or other respiratory complications that may lead to the need for mechanical ventilation. Ventilator duration is a critical factor in the management of patients requiring mechanical ventilation, particularly in the ICU. The use of a ventilator can prolong the lifespan of patients, and the duration of ventilator use may also be a factor that can affect the outcome. During mechanical ventilation, the goal is to maintain adequate gas exchange and ensure that the patient’s blood is fully oxygenated, which includes removing carbon dioxide (CO2) from the body. If the PaCO2 levels are too high (hypercapnia) or too low (hypocapnia), it indicates that the patient is not effectively ventilating, which can lead to respiratory acidosis and potentially life-threatening complications. Monitoring the maximum PaO2 level helps doctors evaluate the oxygenation of blood in a patient’s lungs and whether adjustments to the ventilator settings are necessary to maintain optimal oxygen levels [25]. We also considered patients with various respiratory system diseases specifically in respiratory dysfunction from the MIMIC-III database.

### 2.5. Ablation process

We planned to determine if the currently selected 14 features would negatively impact the model’s performance. We decided to progressively eliminate variables that had a negative effect on the model’s performance, assessing the model’s performance on the validation set by calculating the 95% CI for AUROC. We sequentially removed one variable at a time and assessed the resulting degradation in model performance. The variable that caused the most significant deterioration in model performance was identified and removed in each round. This process was repeated until further removal of variables did not result in a noticeable improvement in model performance. This approach allows us to filter out variables that do not contribute significantly to the predictive power of the model. After applying this iterative feature selection process, we retained 12 out of the 14 initially selected variables. We excluded the variables malignancy and respiratory dysfunction from the final set of features, as they were found to have a negative impact on the model’s performance.

### 2.6. Modeling

The dataset was imbalanced between the number of survivors and non-survivors, with 14,441 survivors and 2,058 non-survivors. The Synthetic Minority Over-sampling Technique (SMOTE) method was used to address the data imbalance issue. Moreover, the train_test_split method was utilized for hierarchical stratified sampling. The dataset was split into three groups: training set, test set and validation set. We proposed a novel deep learning neural network to predict the mortality of ICU patients with mechanical ventilation. Seven baseline ML models were established for result comparison, including KNN, Logistic Regression, Decision Tree, Random Forest, Bagging, XGBoost, SVM[28, 29, 30, 31, 32, 33, 34, 35].

The proposed model is a fully connected neural network comprising an input layer with a dimensionality of 12, followed by a batch normalization (BN) layer for input normalization to improve the stability of the model [36]. Subsequently, three hidden layers are incorporated, each utilizing the rectified linear unit (ReLU) activation function. Between these hidden layers, dropout (DP) layers are employed to randomly discard 50% of the neurons, mitigating overfitting [37]. The first hidden layer consists of 100 neurons, the second hidden layer consists of 50 neurons, and the third hidden layer consists of 25 neurons. The model concludes with an output layer containing a single neuron, utilizing the sigmoid activation function for binary classification with an output probabilities between 0 and 1. This architecture is designed to capture complex patterns in the data while addressing potential overfitting through the strategic use of dropout layers. Figure 2 shows the architecture of our NN model. The model is trained with the Adam optimizer, using binary cross-entropy as the loss function and the AUROC as the evaluation criterion. The training process is run for 100 epochs with a batch size of 256. The model iteratively refines its parameters to minimize the loss function and enhance AUROC performance, aiming to improve its ability to discriminate between positive and negative instances.

**Figure 2:**
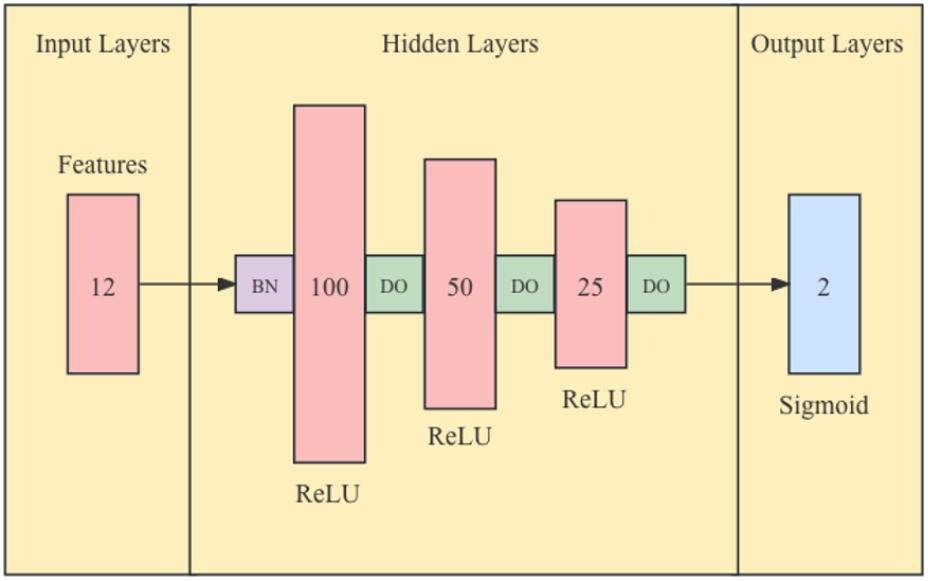
Neural network architecture op-level. This figure shows the details of the neural network architecture.

For the KNN model, we performed Grid Search CV to find the optimal n_neighbors parameter within the range of 1 to 20. The cross-validation (cv) was set to 5, with AUROC employed as the evaluation metric. Regarding the Logistic Regression model, the maximum iteration was set to 1000, and ‘liblinear’ was chosen as the solver. We utilized Grid Search CV across all baseline ML models to identify the best hyperparameter values, optimizing the performance of each prediction model. This process automated the optimization of hyperparameters.

The best model was chosen based on its performance in AUROC on the validation set. Calibration plots were created to assess the models’ accuracy in making probabilistic predictions. A well-calibrated model should have a calibration curve that closely follows the diagonal line, indicating that the predicted probabilities accurately reflect the true likelihood of the outcome. Also, we calculated accuracy for evaluating our models’ performance. The AUROC metric is less affected by class imbalance than accuracy and gives a better picture of the model’s discriminative ability.

In our project, we extracted the dataset using BigQuery, performed data cleaning and conducted model training using Python 3.9.17. The models were derived from the Python libraries scikit-learn 1.2.2 and TensorFlow 2.14.0.

## 3. RESULTS

### 3.1. Cohort Comparison

We obtained 16,499 patients from the MIMIC-III database for model establishment in the patient extraction part. The cohort was then randomly split into a 70% training set, 15% test set, and 15% validation set, respectively, allocating 11,549 patients to the training set, 2,475 patients to the test set, and 2,475 patients to the validation set. The train and validation cohorts were used to train the models. The model with the highest AUROC value was chosen as the best prediction model which was utilized for further assessment on the test set. Table 2 illustrates the comparison of the training set and validation set. In terms of respiratory failure, the proportion of patients (36%) was slightly greater than the validation cohort (35%). The training set (34%) also had slightly higher the share of patients with diabetes than the validation set (33%). We used the Chi-square test to detect whether there is a significant difference in the distribution of categorical features between the training set and the validation set. The null hypothesis (H0) is that the distribution of a particular feature is not significantly different between the training set and the validation set, meaning the feature’s distribution is independent across these sets.However, the p-values of respiratory failure and diabetes were 0.380 and 0.248, respectively, showing no significant difference between cohorts. This suggests that the train and validation cohorts are well-matched, supporting the validity of our model’s training process. Moreover, there was no significant difference among all variables between cohorts based on their p-values.

**Table 2.** Characteristics between train cohort (N=11,549) and validation cohort (N=2,475) with P value. Demographic: Age. Disease severity: SAPS II. Diagnosis: Respiratory dysfunction, Respiratory failure, CHF, Diabetes, Malignancy. Laboratory results: Maximum hemoglobin (g/dl), Minimum lactate (mmol/L), Minimum BUN (mg/dl), Minimum PaO2(mmHg), Maximum PaCO2(mmHg), Minimum PaCO2(mmHg). Others: Vent Duration (Hour).

|  | Train cohort (N=11,549) | Validation cohort (N=2,475) | P |
| --- | --- | --- | --- |
| **Demographic** |  |  |  |
| Age (years) | 63.6(22.0) | 63.6(22.0) | 0.985 |
| **Disease severity** |  |  |  |
| SAPS II | 40.4(19.0) | 40.2(18.0) | 0.515 |
| **Diagnosis** |  |  |  |
| Respiratory dysfunction | 4,273(0.37) | 892(0.36) | 0.382 |
| Respiratory failure | 4,114(0.36) | 858(0.35) | 0.380 |
| CHF | 4,109(0.36) | 888(0.36) | 0.795 |
| Diabetes | 3,908(0.34) | 807(0.33) | 0.248 |
| Malignancy | 1,468(0.13) | 297(0.12) | 0.350 |
| **Laboratory results** |  |  |  |
| Maximum hemoglobin (g/dl) | 12.2(2.6) | 12.2(2.4) | 0.414 |
| Minimum lactate (mmol/L) | 1.3(0.6) | 1.3(0.6) | 0.612 |
| Minimum BUN (mg/dl) | 13.3(8.0) | 13.4(8.0) | 0.653 |
| Minimum PaO<sub>2</sub>(mmHg) | 67.7(45.0) | 68.3(46.0) | 0.531 |
| Maximum PaCO<sub>2</sub>(mmHg) | 58.9(17.0) | 58.0(16.0) | 0.056 |
| Minimum PaCO<sub>2</sub>(mmHg) | 31.1(8.0) | 31.2(8.0) | 0.728 |
| **Treatment** |  |  |  |
| Vent Duration (Hour) | 98.2(102.2) | 97.7(95.1) | 0.900 |
| **Target** |  |  |  |
| Dead in ICU or not | 1,441(0.12) | 307(0.12) | 0.947 |

In addition, a detailed comparison between the survivors group and the non-survivors group was presented in Table 3. Diagnosis and target variables were displayed on the number of diagnosed patients or non-survivors with their proportion, other variables were calculated using the median with standard deviation in parentheses to provide both the central tendency and the variability of the data. The p-values between two subgroups were calculated using the t-test, with the significance level set as P < 0.05. All variables had significant differences between the two groups, indicating a higher association with mortality, except for malignancy.

**Table 3.** Characteristics between survivors (N=14,441) and non-survivors (N=2,058) with P value. Demographic: Age. Disease severity: SAPS II. Diagnosis: Respiratory dysfunction, Respiratory failure, CHF, Diabetes, Malignancy. Laboratory results: Maximum hemoglobin (g/dl), Minimum lactate (mmol/L), Minimum BUN (mg/dl), Minimum PaO2(mmHg), Maximum PaCO2(mmHg), Minimum PaCO2(mmHg). Others: Vent Duration (Hour).

|  | Survivors (N=14,441) | Non-survivors (N=2,058) | P |
| --- | --- | --- | --- |
| **Demographic** |  |  |  |
| Age (years) | 63.2(21.0) | 66.6(22.0) | <0.0001 |
| **Disease severity** |  |  |  |
| SAPS II | 38.6(17.0) | 52.4(23.0) | <0.0001 |
| **Diagnosis** |  |  |  |
| Respiratory dysfunction | 4,918(0.34) | 1,112(0.54) | <0.0001 |
| Respiratory failure | 4,721(0.33) | 1,090(0.53) | <0.0001 |
| CHF | 5,085(0.35) | 789(0.38) | 0.0060 |
| Diabetes | 4,956(0.34) | 600(0.29) | <0.0001 |
| Malignancy | 1,867(0.13) | 251 (0.12) | 0.3715 |
| **Laboratory results** |  |  |  |
| Maximum hemoglobin (g/dl) | 12.3(2.5) | 11.8(2.8) | <0.0001 |
| Minimum lactate (mmol/L) | 1.2(0.5) | 1.9(1.0) | <0.0001 |
| Minimum BUN (mg/dl) | 12.3(8.0) | 19.7(13.8) | <0.0001 |
| Minimum PaO<sub>2</sub>(mmHg) | 69.1(48.0) | 56.6(32.0) | <0.0001 |
| Maximum PaCO<sub>2</sub>(mmHg) | 58.1(15.0) | 62.7(23.0) | <0.0001 |
| Minimum PaCO<sub>2</sub>(mmHg) | 31.4(7.0) | 28.7(8.0) | <0.0001 |
| **Treatment** |  |  |  |
| Vent Duration (Hour) | 89.4(81.7) | 167.2(194.0) | <0.0001 |

### 3.2. Ablation Study on Variable

In our validation set, we evaluated the impact of each variable on the model performance by dropping one variable at a time, measured by AUROC. Firstly, we collected 65 variables from related literature research and applied feature importance to select 14 variables. After training the model, we found that the model with 14 features yielded an AUROC of 0.862. After excluding the ‘malignancy’ variable, the model achieved a slightly increase in AUROC to 0.864, suggesting that ‘malignancy’ may not significantly contribute to the outcome. After removing the ‘respiratory dysfunction’ variable, the AUROC improved to 0.866. This iterative process indicated a potential improvement in model performance. Respiratory failure is a type of disease within the category of ‘respiratory dysfunction’. Retaining respiratory failure while removing respiratory dysfunction indicates that the confirmed diagnosis of respiratory failure has a higher impact on the patient’s survival rate compared to other respiratory dysfunctions. In the end, we obtained the 12 most important variables for model establishment. Figure 3, Figure 4 and Figure 5 show the AUROC improvement process of deleting insignificant features one by one.

**Figure 3:**
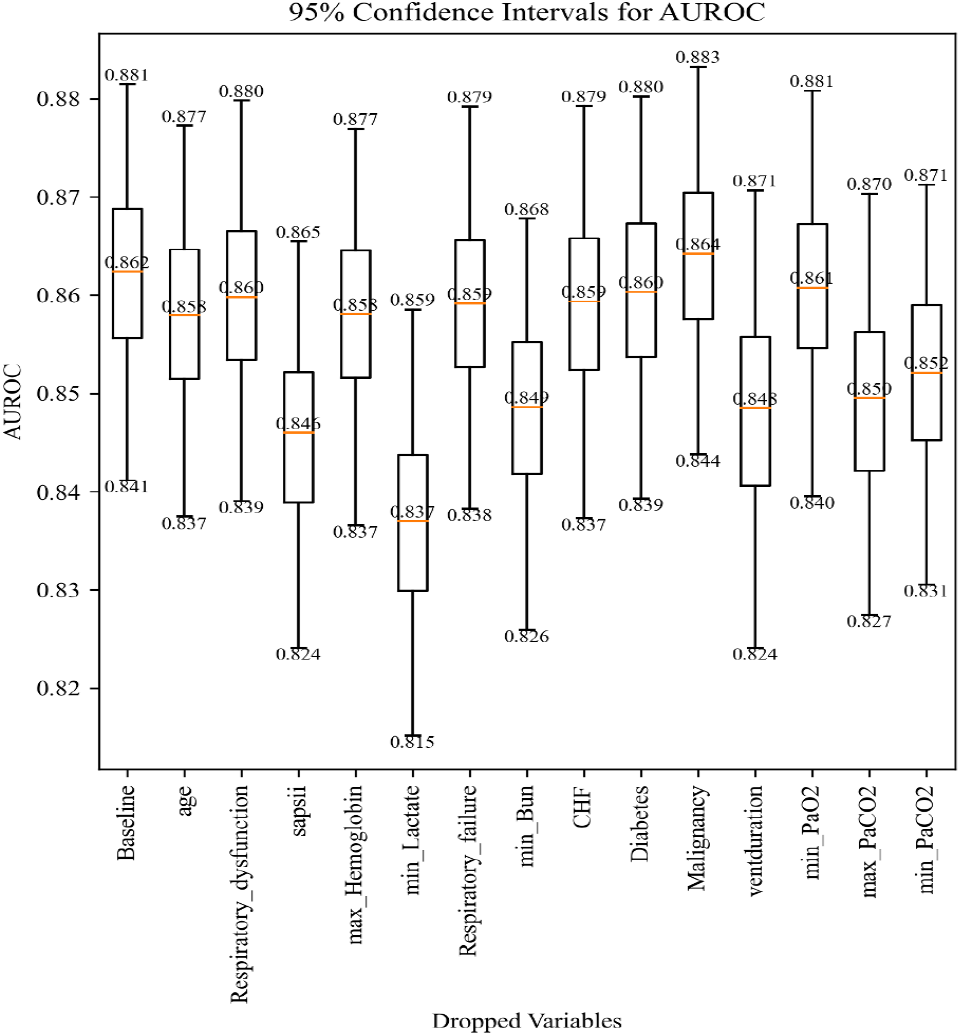
AUROCs boxplots of neural network models with 14 features. Each boxplot displays the AUROC with 95% CI after deleting the corresponding variables. The column baseline shows the result that keeps all the variables. Nothing has been deleted.

**Figure 4:**
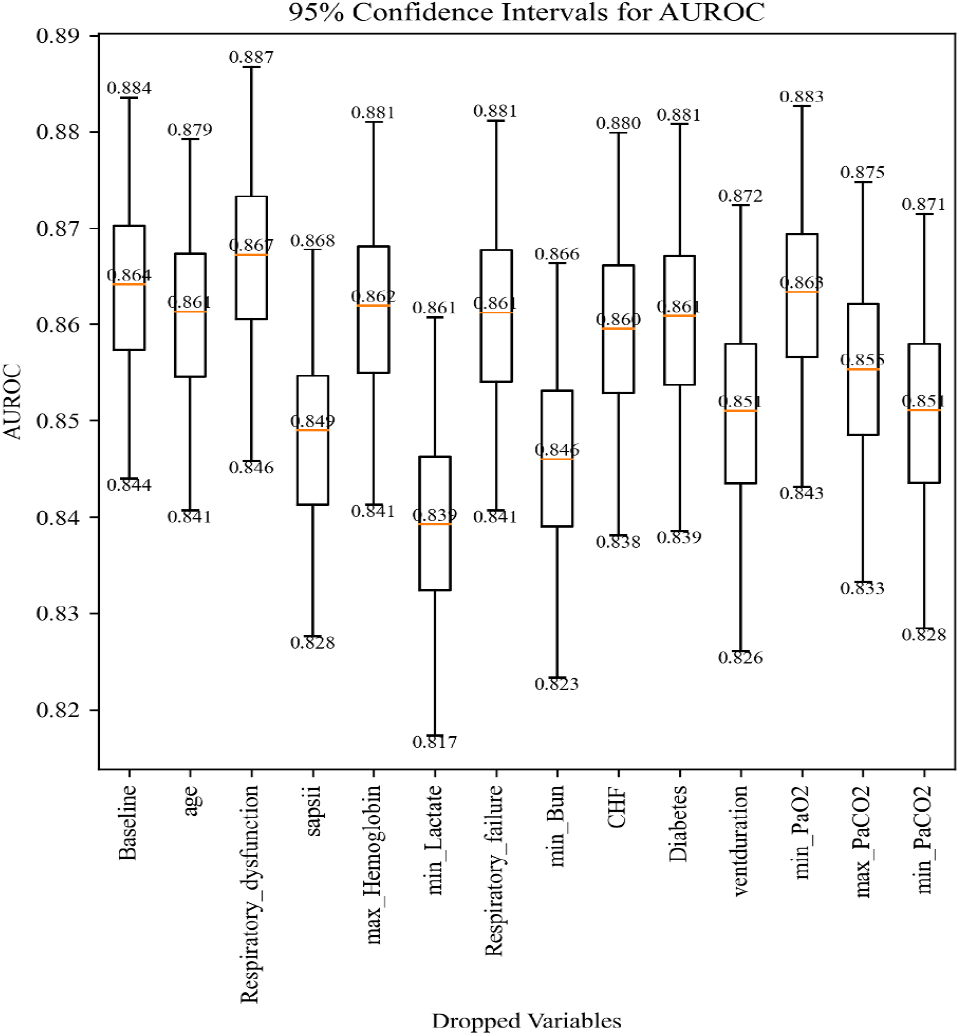
AUROCs boxplots of neural network models with 13 features. Each boxplot displays the AUROC with 95% CI after deleting the corresponding variables. The column baseline shows the result that keeps all the variables. Malignancy has been deleted.

**Figure 5:**
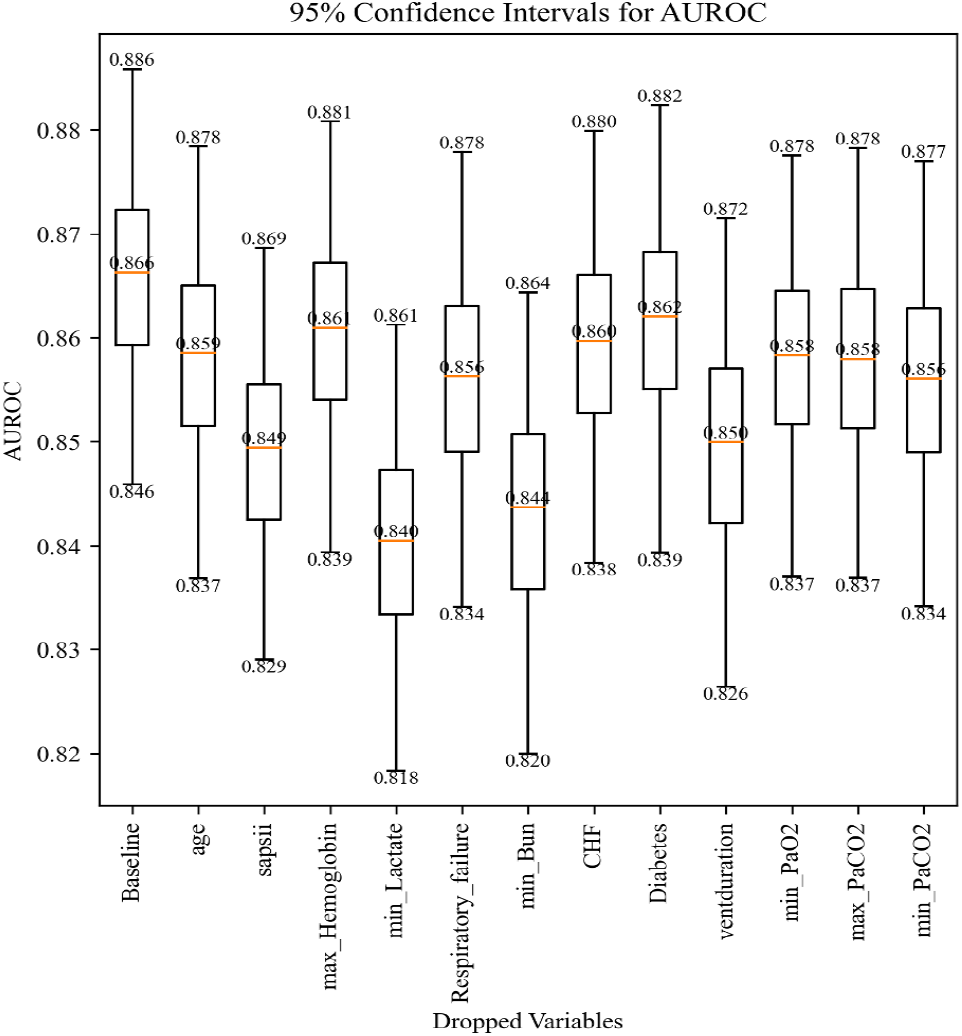
AUROCs boxplots of neural network models with 12 features. Each boxplot displays the AUROC with 95% CI after deleting the corresponding variables. The column baseline shows the result that keeps all the variables. Respiratory dysfunction has been deleted.

### 3.3. Evaluation results

Table 4 and Table 5 show the detailed results summary of our proposed model and baseline ML models. Our proposed neural network model resulted in test set, validation set, and training set, AUROC=0.879, 95% CI = [0.860-0.896], AUROC=0.866, 95% CI = [0.846-0.886], and AUROC=0.958, 95% CI = [0.955-0.960]. The baseline models encompassed KNN, Logistic Regression, Decision Tree, Random Forest, Bagging, XGBoost, and SVM algorithms, yielded the following scores: 0.605, 95% CI [0.578-0.634], 0.851, 95% CI= [0.829-0.871], 0.623, 95% CI = [0.595-0.652]], 0.809, 95% CI = [0.784-0.833], 0.765, 95% CI = [0.734-0.794], 0.854, 95% CI = [0.832-0.872], and 0.851, 95% CI = [0.828-0.874], respectively. We also calculated the accuracy score of KNN 0.809, Logistic Regression 0.783, Decision Tree 0.809,Random Forest 0.860, Neural Network 0.859, Bagging 0.845, XGBoost 0.878, and SVM 0.881. These scores provided insights into how well the models would perform on new unseen data.

**Table 4.** Evaluation results and confidence interval for proposed model. Trained the model on different sets: test set, validation set and training set. The evaluation metrics included AUROC, AUROC 95% CI, precision, recall value, accuracy score, and F1 score.

| Models | AUROC | AUROC 95% CI | Accuracy |
| --- | --- | --- | --- |
| Proposed model performance on test set | 0.879 | [0.860-0.896] | 0.859 |
| Proposed model performance on validation set | 0.866 | [0.846-0.886] | 0.855 |
| Proposed model performance on training set | 0.958 | [0.955-0.960] | 0.881 |

**Table 5.** Evaluation metrics and confidence interval for seven models. We used a total of seven different models, KNN, Logistic Regression, Decision Tree, Random Forest, Bagging, XGBoost, and SVM. The evaluation metrics included AUROC, AUROC 95% CI and accuracy score.

| Models | AUROC | AUROC 95% CI | Accuracy |
| --- | --- | --- | --- |
| KNN | 0.605 | [0.578-0.634] | 0.809 |
| Logistic Regression | 0.851 | [0.829-0.871] | 0.783 |
| Decision Tree | 0.623 | [0.595-0.652] | 0.809 |
| RF | 0.809 | [0.784-0.833] | 0.860 |
| Bagging | 0.765 | [0.734-0.794] | 0.845 |
| XGBoost | 0.854 | [0.832-0.872] | 0.876 |
| SVM | 0.851 | [0.828-0.874] | 0.881 |

Figure 6 and Figure 7 display Receiver Operating Characteristic (ROC) curves of our proposed model neural network and the seven baseline models, KNN, Logistic Regression, Decision Tree, Random Forest, Bagging, XGBoost, and SVM on test set and validation set. We observed that all models except KNN and Logistic Regression exhibited smoother ROC curves and achieved higher AUROC values. Figure 8 displays the AUROC boxplots of our proposed model and baseline models. We observed that the AUROCs of Logistic Regression, neural network, XGBoost, and SVM exceeding 0.8, showing the strong predictive ability. Among these boxplots, the mean AUROC of our proposed model exceeds the maximum value of the AUROCs of other baseline models.

**Figure 6:**
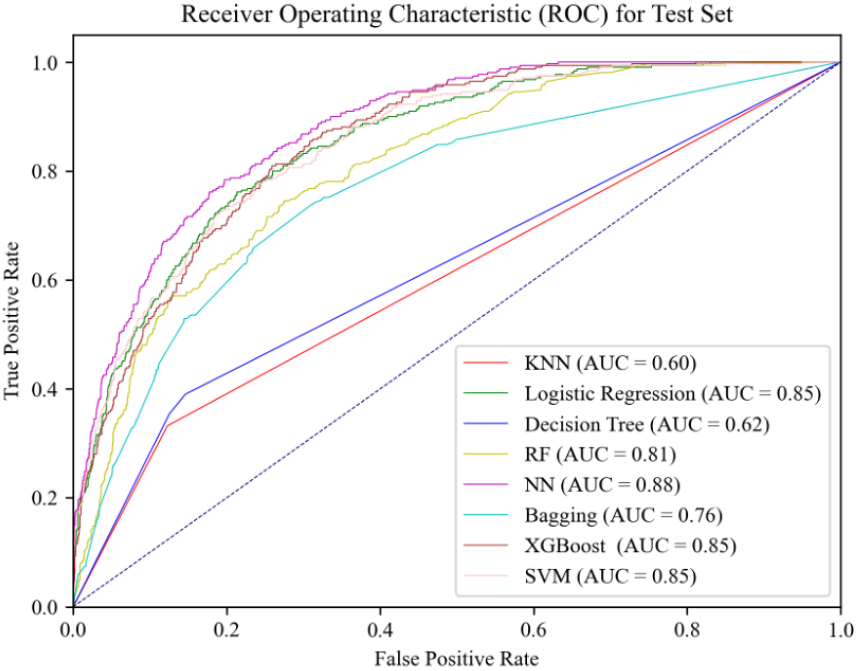
ROC curves of the eight models for the test set. KNN, Logistic Regression, Decision Tree, Random Forest, Neural Network, Bagging, XGBoost, and SVM.

**Figure 7:**
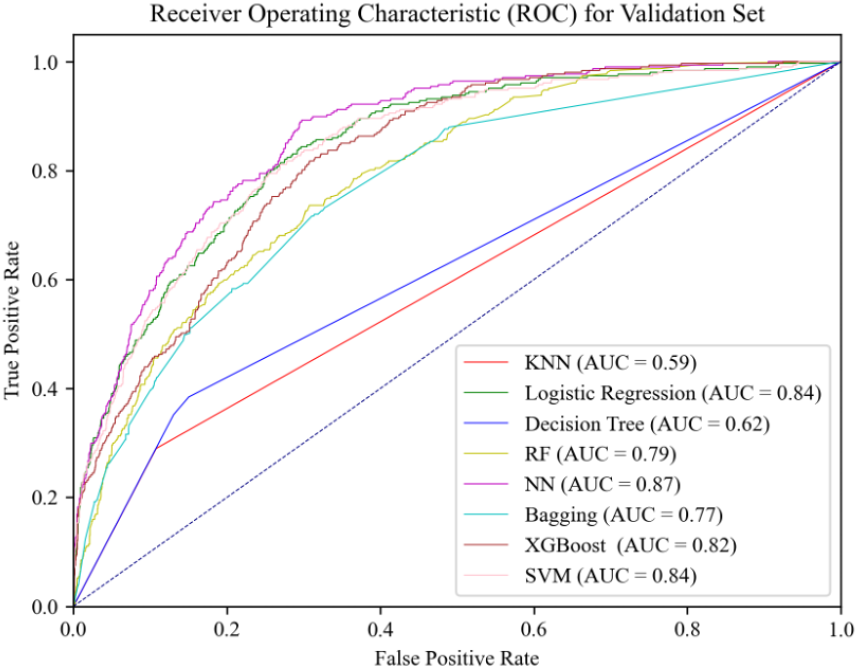
ROC curves of the eight models for the validation set. KNN, Logistic Regression, Decision Tree, Random Forest, Neural Network, Bagging, XGBoost, and SVM.

**Figure 8:**
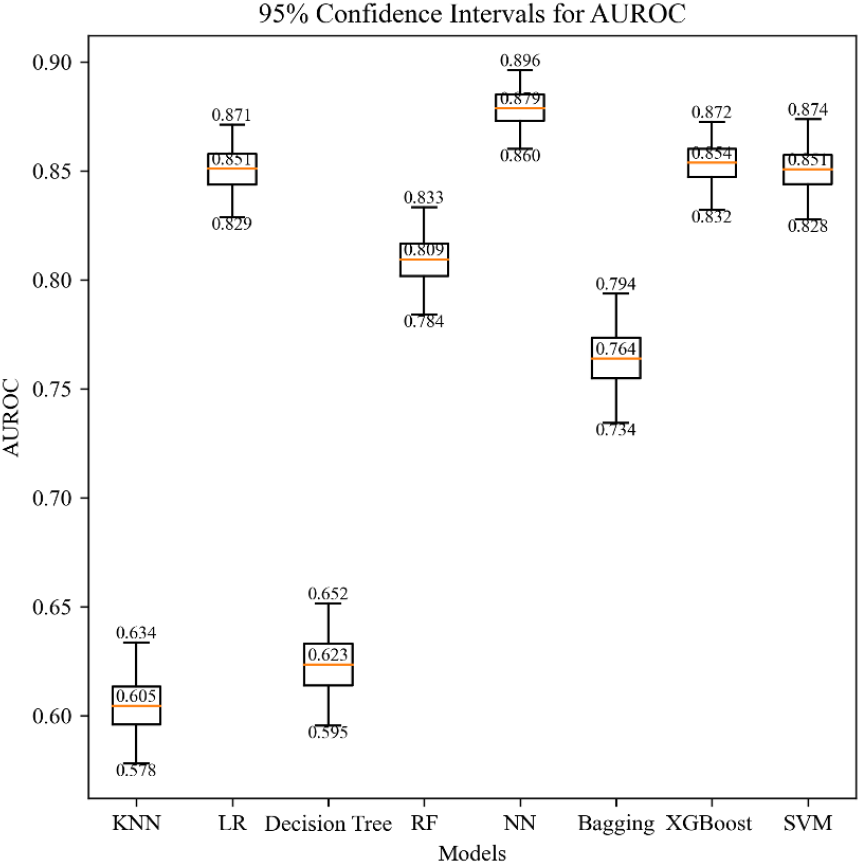
AUROCs boxplots of the eight models. KNN, Logistic Regression, Decision Tree, Random Forest, Neural Network, Bagging, XGBoost, and SVM. The upper line of the single boxplot represents the maximum value of AUROC, the lower line represents the minimum value of AUROC, and the middle line represents the mean value of AUROC.

Also, we applied calibration techniques and rigorous evaluation methods. Calibration plots were generated by plotting the mean predicted probability against the observed frequency of outcomes in each decile. The Brier Score measured the mean squared difference between predicted probabilities and actual outcomes, with lower scores indicating better calibration. Isotonic Regression, a non-parametric method, was used for calibration. Figure 9 shows that the predicted probabilities are well-calibrated, with points close to the diagonal line. Among the models, the two best-performing ones in terms of Brier score were SVM (0.0905) and neural network (0.0974), demonstrating very good calibration. These results confirm that our model’s predicted probabilities are highly accurate and well-calibrated. The low Brier Score and high AUROC substantiate its accuracy and reliability. These findings align with existing literature on well-calibrated prediction models and demonstrate the strength of our approach in providing reliable predictions for clinical decision-making in the ICU setting.

**Figure 9:**
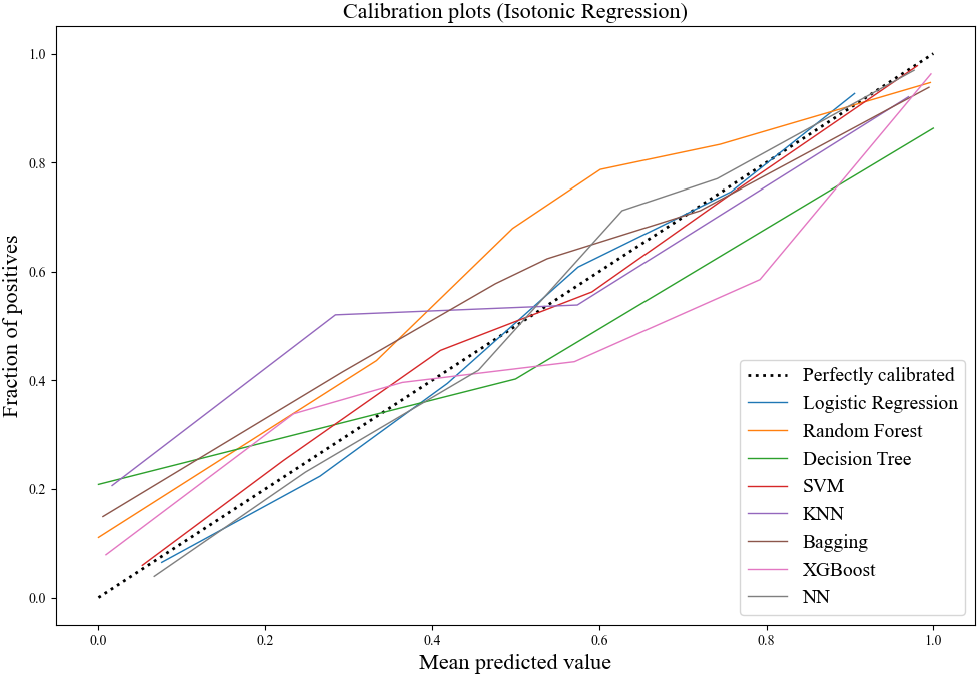
Calibration plots of the eight models. KNN, Logistic Regression, Decision Tree, Random Forest, Neural Network, Bagging, XGBoost, SVM.

### 3.4. SHAP analysis

SHAP (SHapley Additive exPlanations) is a method used in ML to understand the impact of individual variables on model predictions. It provides a way to interpret the output of any ML model by quantifying the contribution of each feature to the predicted outcome [38]. Figure 10, along with Table 6, displays the SHAP (SHapley Additive exPlanations) values for the test set, providing a detailed analysis of how each variable affects the model’s prediction. The SHAP analysis identifies ‘respiratory failure’ as the most significant predictor, followed by ‘minimum lactate’, ‘SAPS II’, ‘ventilation duration’, ‘minimum BUN’, ‘diabetes’, ‘maximum PaCO2’, ‘CHF’, ‘minimum PaCO2’, ‘age’, ‘minimum PaO2’ and ‘maximum hemoglobin’. Here, the ‘respiratory failure’ shows a notable positive impact on the model’s predictions. The ‘minimum PaO2’ and ‘maximum hemoglobin’, in comparison, demonstrate more moderate effects. The shift in the order of feature importance in the training set highlights the importance of considering diverse metrics for a comprehensive model evaluation.

**Table 6.** Average SHAP value for 12 features in the test set. The 12 predictors: Respiratory failure, diabetes, age, SAPS II Score, maximum hemoglobin, minimum lactate, CHF, vent duration, minimum bun, minimum PaCO2, maximum PaCO2, minimum PaO2.

| Features | Average SHAP Value (Test Set) |
| --- | --- |
| Respiratory Failure | 0.096 |
| Minimum Lactate | 0.080 |
| SAPS II | 0.075 |
| Vent Duration | 0.075 |
| Minimum Bun | 0.052 |
| Diabetes | 0.043 |
| Maximum PaCO<sub>2</sub> | 0.040 |
| CHF | 0.034 |
| Minimum PaCO<sub>2</sub> | 0.025 |
| Age | 0.019 |
| Minimum PaO<sub>2</sub> | 0.010 |
| Maximum Hemoglobin | 0.008 |

**Figure 10:**
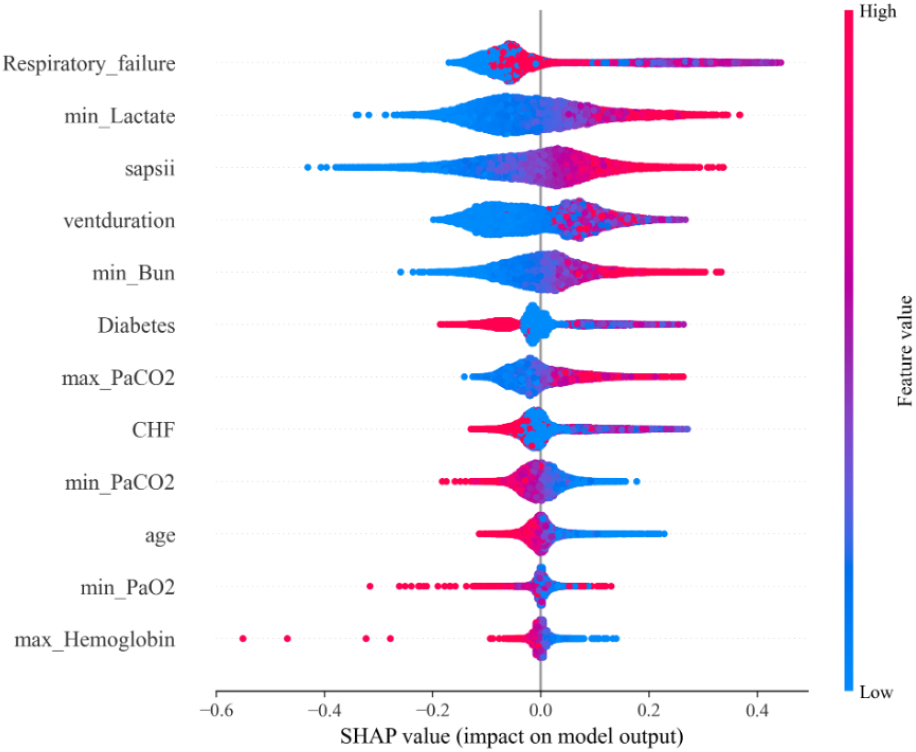
SHAP value based on neural network model for the test set. Predictors: Respiratory failure, diabetes, age, SAPS II Score, maximum hemoglobin, minimum lactate, CHF, vent duration, minimum bun, minimum PaCO2, maximum PaCO2, minimum PaO2.

The difference between the feature importance ranking in Figure 11 and the average SHAP value in Table 6 could be attributed to the different methodologies underlying these two approaches. It is also worth mentioning that while ‘CHF’ and ‘diabetes’ held moderate importance in the SHAP ranking, they appeared as top contributors in the feature importance ranking. This divergence underscores the complexity of variable interactions within the model and highlights the necessity of employing multiple interpretability methods to fully understand the model’s behavior.

**Figure 11:**
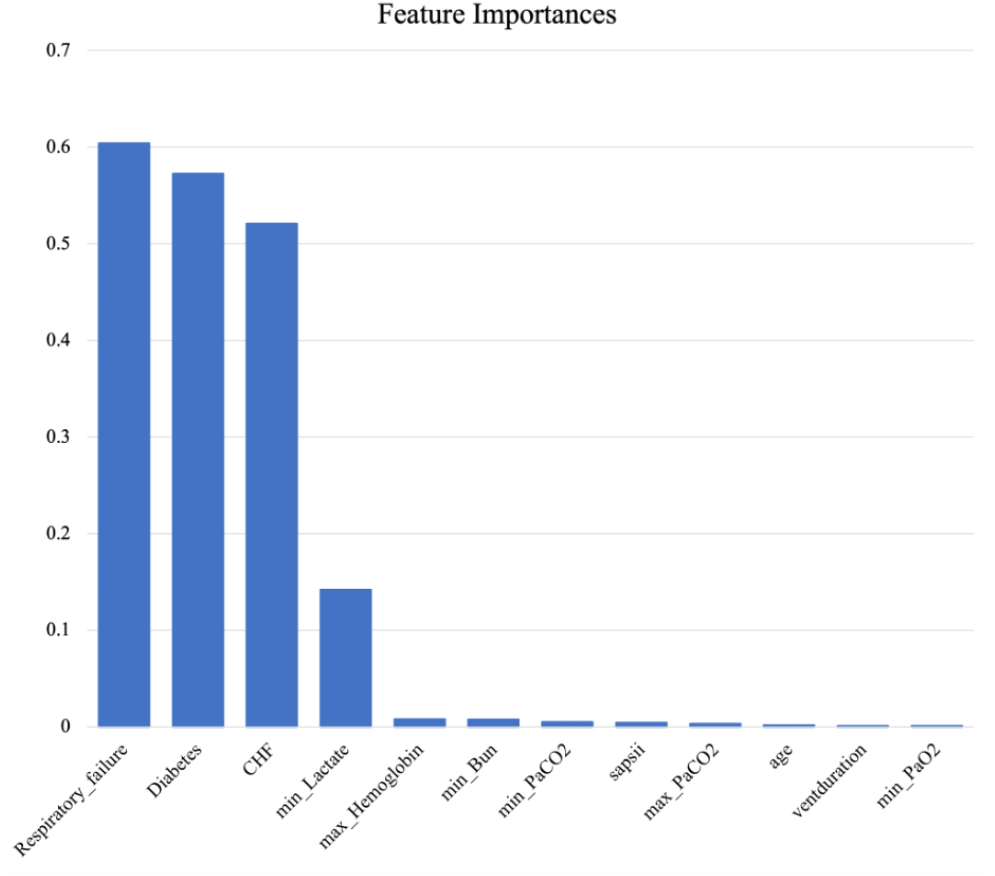
Feature importance is based on Neural Network model for the test set. Predictors: Respiratory failure, diabetes, age, SAPS II Score, maximum hemoglobin, minimum lactate, CHF, vent duration, minimum bun, minimum PaCO2, maximum PaCO2, minimum PaO2.

Furthermore, the minimal impact of ‘minimum PaCO2’ and ‘maximum Hemoglobin’ on the model output, as indicated by their low feature importance and SHAP values, suggests that these factors are less discriminative for the predictive task at hand. The comprehensive analysis of these indicators provides valuable insights into the model’s decision-making process, guiding practitioners in refining the model and focusing on the most pertinent predictors for outcome prediction.

## 4. DISCUSSION

### 4.1. Existing model compilation summary

In our study, we proposed a neural network model to predict the ICU mortality of patients undergoing invasive mechanical ventilation. The result of our mortality prediction model was better than the best existing literature by Y.Zhu et al [19]. The effectiveness of our model demonstrated 7.06% improvement in AUROC.

Although the existing literature result effectively predicted mortality rates among ICU patients, it exhibited certain limitations. They used a total of 66 variables to predict the model outcome. This approach may raise concerns related to model complexity and overfitting. Additionally, the outcome of their research was considered unsatisfactory and inadequate for practical use in clinical exercising.

For our research, we used advanced feature selection techniques to select only 12 variables as our features. The results of our model had a significant improvement, which provided a more reliable result for clinical use and highlights the efficiency and effectiveness of our model in delivering superior predictive performance with a more concise feature set. Moreover, we used the training set, validation set and test set for evaluation. The best existing model only used training and test sets. Using different sets for hyperparameter tuning and model evaluation could avoid information leakage and enhance the model’s generalization assessment capabilities. Furthermore, our proposed neural network model was easy set up and replicated, had fewer layers which helps prevent overfitting and trains faster than other deep neural networks.

These features made our model efficient and practical for real-world applications.

We found that respiratory failure had a higher association with patient mortality than respiratory dysfunction from SHAP analysis and feature importance, which means the condition of respiratory failure was much more important than the general comprehensive disease of respiratory dysfunction. This suggests that clinicians should pay more attention to patients with a history of respiratory failure. The ventilation duration was also a clinically meaningful variable that directly reflects the severity and duration of respiratory failure. It provides insight into the patient’s respiratory status and the level of support needed. From the SHAP value analysis (Figure 10), we found that the longer the ventilation use time, the higher the patient’s mortality rate, proving that prolonged use of a ventilator does not improve patient survival.

Additionally, the SVM and XGBoost baseline models had AUROCs of 0.841 and 0.825, respectively, which were similar to the performance of our proposed NN model. We observed that an overlap in the AUROC boxplots of these two models in Figure 8, which illustrates the performances of these two models were also powerful in predicting mortality since NN had multiple layers and required more time for training.

### 4.2. Study limitations

In our model development, we used training and validation datasets to construct the model. The test dataset was used for evaluating the performance of the model. The training, validation, and test sets were all from the MIMIC-III database. However, using independent datasets from different systems would be beneficial for testing the performance of the model. The MIMIC-III database is a large but outdated database, which contains the dataset of related ICU patients only between 2001 and 2012. Exploring newer datasets could enhance the predictive capabilities of our model. Additionally, integrating other types of data, such as images and text, could further improve the accuracy and utility of our results.

## 5. CONCLUSION

The goal of our paper is to build a novel neural network model to predict the mortality of ICU patients undergoing mechanical ventilation. Compared with the results of baseline ML models and existing literature, deep learning methods for modeling ICU patient data in the MIMIC-III database to predict mechanical ventilation mortality have provided significant improvements in predicting observed outcomes. This improvement may be due to the efficiency of the variables, such as the time series variables we selected for predicting the model. Our framework provides valuable support for clinicians to identify patients at high risk of death in the ICU. This predictive tool is particularly beneficial for patients and clinicians, as it can assess the time when a patient leaves the ICU and guide clinicians in arranging patient treatment plans.

Future research could focus on validating our methods with datasets from different healthcare systems or exploring their applicability to various diseases and outcomes. Exploring the applicability of our methods to various diseases and outcomes can uncover new insights and potential applications. Additionally, other researchers could explore different types of data based on our dataset and this can lead to innovative research directions. For example, integrating image data or text data with our existing dataset could provide a more comprehensive understanding of patient health and improve the accuracy of our predictions.

## Data Availability

All data produced in the present study are available upon reasonable request to the authors.

## Acknowledgment

The authors extend their gratitude to the creators of MIMIC-III for furnishing a thorough and inclusive public electronic health record (EHR) dataset [39].

